# The performance of the SARS-CoV-2 RT-PCR test as a tool for detecting SARS-CoV-2 infection in the population: A survey of routine laboratory RT-PCR test results from the region of Münster, Germany

**DOI:** 10.1101/2021.05.06.21256289

**Authors:** Andreas Stang, Johannes Robers, Birte Schonert, Karl-Heinz Jöckel, Angela Spelsberg, Ulrich Keil, Paul Cullen

## Abstract

**Objectives:** To evaluate the population-based performance of the SARS-CoV-2 RT-PCR test as a tool for detecting SARS-CoV-2 infection during the pandemic in 2020.

**Methods:** We analysed SARS-CoV-2 RT-PCR results of 162,457 people living in Münster, Germany screened at nursing homes, testing sites, at schools, regional hospitals, and by general practitioners. All PCRs were done with the same cobas SARS-CoV-2 RT-PCR system (Roche Diagnostics). We stratified positive RT-PCR results by cycle threshold (Ct) values, periods of the national test strategy, age, sex, and symptoms.

**Results:** Among 162,457 individuals, 4164 (2.6%) had a positive RT-PCR test result, defined as Ct<40. Depending on the national test strategy, higher positive rates were associated with testing predominantly symptomatic people. Children (0-9 years) and older adults (70+ years). Only 40.6% of test positives showed low Ct values < 25 (potentially infectious). The percentage of Ct values below 25 was lower among children (0-9), adolescents (10-19), and among the elderly (70+ years).

**Conclusions:** RT-PCR testing as a tool for mass screening should not be used alone as a base for pandemic decision making including measures such as quarantine, isolation, and lockdown.

## Introduction

Worldwide detection and monitoring of SARS CoV-2 infection are based on results of the real-time reverse-transcription polymerase chain reaction (RT-PCR) test, interpreting a positive test as proof of infection with the SARS-CoV-2 virus. This is the first time that pandemic decision making and management relies almost exclusively on PCR testing. In Germany, the national testing strategy and its financial compensation changed several times. During weeks 10-19, 2020, mostly symptomatic people were tested, due to limited testing capacities. After May 2020, testing capacities were hugely expanded to a weekly performance of 1.6 million RT-PCR tests. Between weeks 20-44, RT-PCR testing was performed extensively in asymptomatic population groups, e.g. school teachers, nursing home residents and their caregivers, other health care professionals, meat plant workers and contact persons of RT-PCR test positive individuals. During weeks 28-35, RT-PCR testing focused mainly on travel hubs such as airports, train stations and motorway service areas. After November 3, 2020, a government decree restricted RT-PCR testing to symptomatic persons and recommended the use of rapid diagnostic antigen tests for asymptomatic people.

Between February 27, 2020 and February 9, 2021, more than 2.4 million positive test results out of a total of 42 million tests were counted as incident SARS-CoV-2 infections in Germany [1]. However, a positive RT-PCR test “does not consistently equate to presence of infectious virus” [2, 3] and therefore should not be regarded as a straightforward tool for pandemic planning. A recent systematic-review of infectious potential of RT-PCR test positives showed that patient characteristics (date of onset and severity of symptoms) and test factors (cycle threshold) influence the reliability of the test [4]. Therefore, important questions about the interpretation of a positive RT-PCR test remain to be answered. “RT-PCR tests do not detect the virus, they detect the presence of known genetic sequences from which inferences are drawn” [5].

On January 21, 2021, the WHO declared that careful interpretation of a weak positive RT-PCR test is needed. Where test results including the cycle threshold (Ct) “do not correspond with the clinical presentation, a new specimen should be taken and retested using the same or different PCR technology”. As no standardization for Ct values exists across RT-PCR platforms, it is difficult to compare Ct values derived from different assays using different primers. We present real world data from a large laboratory in Münster, north western Germany, derived from a single fully automated high throughput RT-PCR platform with the same gene targets (E-gene, ORF1a/b gene). Our study aims to describe the changes in the prevalence of positive RT-PCR tests over time, assessed with a consistent methodology during the complete study period. In addition, the influence of covariates such as age, sex, calendar time, and symptoms at the time of first RT-PCR test on the distribution of Ct values and their consistency are analysed and compared with the findings of other Ct value surveys.

## Material and Methods

All test results were obtained from a single laboratory located in the city of Münster (population 313,000), using the cobas SARS-CoV-2 RT-PCR system (Roche Diagnostics). Swab specimens were sent for analysis by regional hospitals, nursing homes, testing sites run by the municipalities, and general practitioners of the region between calendar weeks 10 and 49, 2020. The Münster region covers an area of about 6000 square kilometres located in the north-west of Germany with about 1.64 million inhabitants, the great majority of caucasian ethnicity. Life expectancy at birth is 81.2 years (78.9 for men and 83.5 for women) as of 2017, figures almost identical to those of Germany as a whole. In the Münster region, the cumulative COVID-19 death toll as of February 11, 2021, of 66/100,000 [6], is very similar to that of the entire country [7].

About 80% of all SARS-CoV-2 RT-PCR tests in the Münster region were carried out in our laboratory. During weeks 10-12, about 7000 tests were performed each week in all of Germany, about 500 of which in our laboratory alone. After week 20, testing capacities were massively expanded in Germany, with 196 laboratories performing an average of about 10,000 tests per week each. Nearly all swab specimens in our laboratory were tested within 24 hours of collection. In isolated cases, testing was delayed for up to 72 hours. The tests and their interpretation were carried out in accordance with the Roche cobas SARS-CoV-2 emergency use authorization (EUA) protocol, the specific targets of the test being the open reading frame (ORF) 1ab and the pan-Sarbecovirus E genes. The limit of detection (LoD), defined as the concentration of analyte that will be detected in 95% of replicate tests was 0.007 median tissue culture infectious doses (TCID50) per ml for target 1 and 0.004 TCID50/ml for target 2, corresponding to Ct values of approximately 33 and 36, respectively (cobas® SARS-CoV-2 package insert, version 1.0). RT-PCR tests that had not crossed the positivity threshold after the 40th cycle were reported as “negative”. The Ct value is inversely proportional to the initial amount of target nucleic acid and is thus a relative indicator of the concentration of viral particles in the clinical specimen. Each increase in Ct value of three points corresponds to a reduction in the amount of initial viral material by a factor of about ten.

We extracted RT-PCR test results from the period from March 26 to December 6, 2020 (calendar weeks 12-49). Ct values for positive RT-PCR tests were available for tests performed during calendar weeks 14-15 and 26-49. For technical reasons, we were unable to extract the Ct values for the remaining weeks from the proprietary format of the archives stored on the Roche cobas device. According to the professional code of conduct for physicians of the North Rhine Medical Association, an ethics vote is not required for the performance of a purely retrospective epidemiological study.

## Statistical methods

After exclusion of 232 tests that were uninterpretable and one test with an implausible Ct value, the final data set included 193,258 RT-PCR test results. Two strategies of analysis were pursued: 1. person-based strategy, meaning that only the first test of each person was selected; 2. case-based strategy, meaning that all test results were selected regardless of number of tests performed per individual. The main analyses presented in this paper are based on person-based data. Case-based analyses are shown in the **supplementary files**.

As population data on the distribution of Ct values and the negative correlation between Ct values as an indicator of viral load and infectivity are sparse, we categorized Ct values according to the UK Office for National Statistics (ONS) COVID-19 household survey which showed that

> “people with a higher concentration of viral genetic material (positive cases with low Ct values; below 25) are more likely to be infectious in a household than those with lower concentrations (positive cases with high Ct values; above 25) high Ct values cluster with negative cases – indicating transmission has not been observed.” [8]

As there is some uncertainty about the best cutoff for Ct values, we also categorized Ct values based on a cutoff of < 30 versus ≥ 30. We stratified RT-PCR test results and available Ct values according to the different phases of the national SARS-CoV-2 testing strategy (weeks 10-19, 20-44, and 45-49). In addition, we identified three one week periods in each phase most strongly representing the characteristics of the respective national strategy with the first peak in calendar weeks 12-13 (proxy: weeks 13-14), unselective testing with a peak of test positives among returning vacationers in weeks 33-34, and the third peak in calendar weeks 50-51 (proxy: weeks 48-49).

To compare the Ct value distribution of the population of Münster with the population-based data of the ONS Coronavirus (COVID-19) Infection Survey [9], we analysed the Ct value distributions for corresponding time periods. As a caveat, the UK survey used the Taqpath platform and tested for ORF1ab, N, and S, limiting the ability to match individual Ct values (as opposed to value distributions).

When sending swab tubes for RT-PCR testing, senders were able to note the symptoms of the test subject on the request form. Most of this information was imprecise, so that reliable classification of symptomatic and asymptomatic individuals was possible only in a small subsample. Classification was performed using rule-based text mining, which searched for key words indicating the presence or absence of symptoms of Covid-19 on the basis of a word list. Identification of symptomatic and asymptomatic individuals by text mining was double-checked by referring to the original request form. All analyses were performed using SAS for windows, 9.4. (SAS Institute Inc., Cary, NC, USA). For the text mining, we used the SAS INDEX function of SAS.

## Results

Among 162,457 subjects, a total of 193,258 SARS-CoV-2 RT-PCR tests were performed during the study period, 21,537 (13.3%) had two tests, and 9,264 (5,7%) received more than two tests. Analysing 162,457 first swab specimens, 4,164 individuals (2.6%) had a positive RT-PCR test. Men had a slightly higher positivity rate than women (2.8% versus 2.4%, respectively). The positive rate was also slightly higher if the reported swab sites included nose and throat as opposed to throat only. The percentage of positives was lower among children aged 0-9 years and among adults aged 70 or more, compared to age groups 10-69 years (2.2% and 1.6%, respectively versus 2.8%). The positive rate was also linked to the national SARS-CoV-2 test strategy. During the first and third phase of national testing, corresponding to the flu season, predominantly symptomatic people were tested. During these phases, the positive rates were higher than during the intermittent second phase corresponding to the summer season, when predominantly asymptomatic individuals were tested. The positive rate during the third phase was considerably higher than during the first phase. A similar, albeit greater, difference was found when we stratified our analyses according to the three predefined two-week periods. During the peak of testing asymptomatic individuals, only 0.4% tested positive with a mean Ct value of 28.8. Similarly, higher mean Ct values were observed among children aged 0-9 years and adults above 70 years. Only 40.6% of test positives showed low Ct values (below 25, i.e. more likely to be infectious) (**Table 1, Supplementary Table 1)**.

**Table 1.** Characteristics of people who underwent PCR testing in the region of Münster, North Rhine-Westphalia, Germany, March 26 - December 6, 2020.

|  |  | Number of tests<sup>1)</sup> | Positive tests |  | Mean Ct value among positive tests<sup>2)</sup> |  | Percentage of positive tests with Ct values<sup>2)</sup> |  |
| --- | --- | --- | --- | --- | --- | --- | --- | --- |
|  |  | N | N | % | Mean | SD | < 25 | <30 |
| All |  | 162,457 | 4164 | 2.6 | 26.5 | 5.2 | 40.6 | 69.6 |
|  | Men | 70,043 | 1981 | 2.8 | 26.4 | 5.3 | 42.0 | 69.6 |
|  | Women | 92,113 | 2165 | 2.4 | 26.6 | 5.1 | 39.4 | 69.5 |
|  | Unknown | 301 | 18 | 6.0 | 27.4 | 5.2 | 38.9 | 66.7 |
| Swab site |  |  |  |  |  |  |  |  |
|  | Nose & throat | 8637 | 222 | 2.6 | 25.9 | 5.4 | 43.0 | 72.9 |
|  | Throat | 7059 | 151 | 2.1 | 26.2 | 4.5 | 41.7 | 77.2 |
|  | Unspecified/oth | 146,761 | 3791 | 2.6 | 26.6 | 5.2 | 40.4 | 69.1 |
| er |  |  |  |  |  |  |  |  |
| Age group |  |  |  |  |  |  |  |  |
|  | 0-9 | 9978 | 222 | 2.2 | 28.6 | 4.7 | 21.1 | 56.5 |
|  | 10-19 | 15,200 | 536 | 3.5 | 26.8 | 4.9 | 38.2 | 71.4 |
|  | 20-29 | 21,613 | 745 | 3.5 | 26.4 | 5.1 | 41.6 | 69.4 |
|  | 30-39 | 21,830 | 572 | 2.6 | 26.3 | 5.1 | 42.7 | 72.3 |
|  | 40-49 | 21,373 | 600 | 2.8 | 26.3 | 5.4 | 43.8 | 69.1 |
|  | 50-59 | 25,367 | 665 | 2.6 | 26.0 | 5.3 | 44.4 | 72.9 |
|  | 60-69 | 17,460 | 351 | 2.0 | 26.0 | 5.1 | 46.0 | 73.5 |
|  | 70-79 | 12,155 | 214 | 1.8 | 27.1 | 5.2 | 35.3 | 65.8 |
|  | 80-89 | 13,196 | 185 | 1.4 | 26.8 | 5.2 | 37.4 | 64.5 |
|  | 90-99 | 3699 | 55 | 1.5 | 27.0 | 5.4 | 37.0 | 63.0 |
|  | 100+ | 29 | 1 |  |  |  |  |  |
|  | unknown | 557 | 18 | 3.2 | 31.3 | 4.9 | 11.8 | 29.4 |
| Calendar week |  |  |  |  |  |  |  |  |
|  | 10-19 | 12,985 | 305 | 2.4 | 28.7 | 5.1 | 22.1 | 46.8 |
|  | 20-44 | 132,488 | 2418 | 1.8 | 26.5 | 5.2 | 40.5 | 69.6 |
|  | 45-49 | 16,984 | 1441 | 8.5 | 26.4 | 5.1 | 41.8 | 70.7 |
| Specific phases of the pandemic<sup>3)</sup> |  |  |  |  |  |  |  |  |
|  | Peak 1<sup>st</sup> wave | 2190 | 36 | 1.6 | 27.8 | 5.4 | 26.5 | 55.9 |
|  | Traveler return | 16,874 | 68 | 0.4 | 28.8 | 5.5 | 26.9 | 55.2 |
|  | Peak 2<sup>nd</sup> wave | 4022 | 367 | 9.1 | 26.6 | 5.1 | 39.5 | 69.8 |

Legend table: SD = standard deviation; <sup>1)</sup> only persons with tests that were clearly either positive or negative were included; <sup>2)</sup> among 4164 people tested positive, the Ct value was available for 3810 people (91.5%); Ct values were not retrievable for positive tests during the calendar weeks 12-13 and 16-25 in 2020; <sup>3)</sup> Peak of 1<sup>st</sup> wave in weeks 12-13 (16.-29.3.2020); proxy weeks 13-14; unselective testing in weeks 33-34 (peak of tests for traveler return); peak of 2<sup>nd</sup> wave in weeks 50-51 (7.-20.12.2020), proxy weeks 48-49

Among RT-PCR test positives, 19 asymptomatic and 39 symptomatic individuals were identified by text mining. **Figure 1** shows that mean Ct values among symptomatic subjects were markedly lower than among asymptomatic individuals (25.5 vs 29.6, respectively).

**Figure 1.**
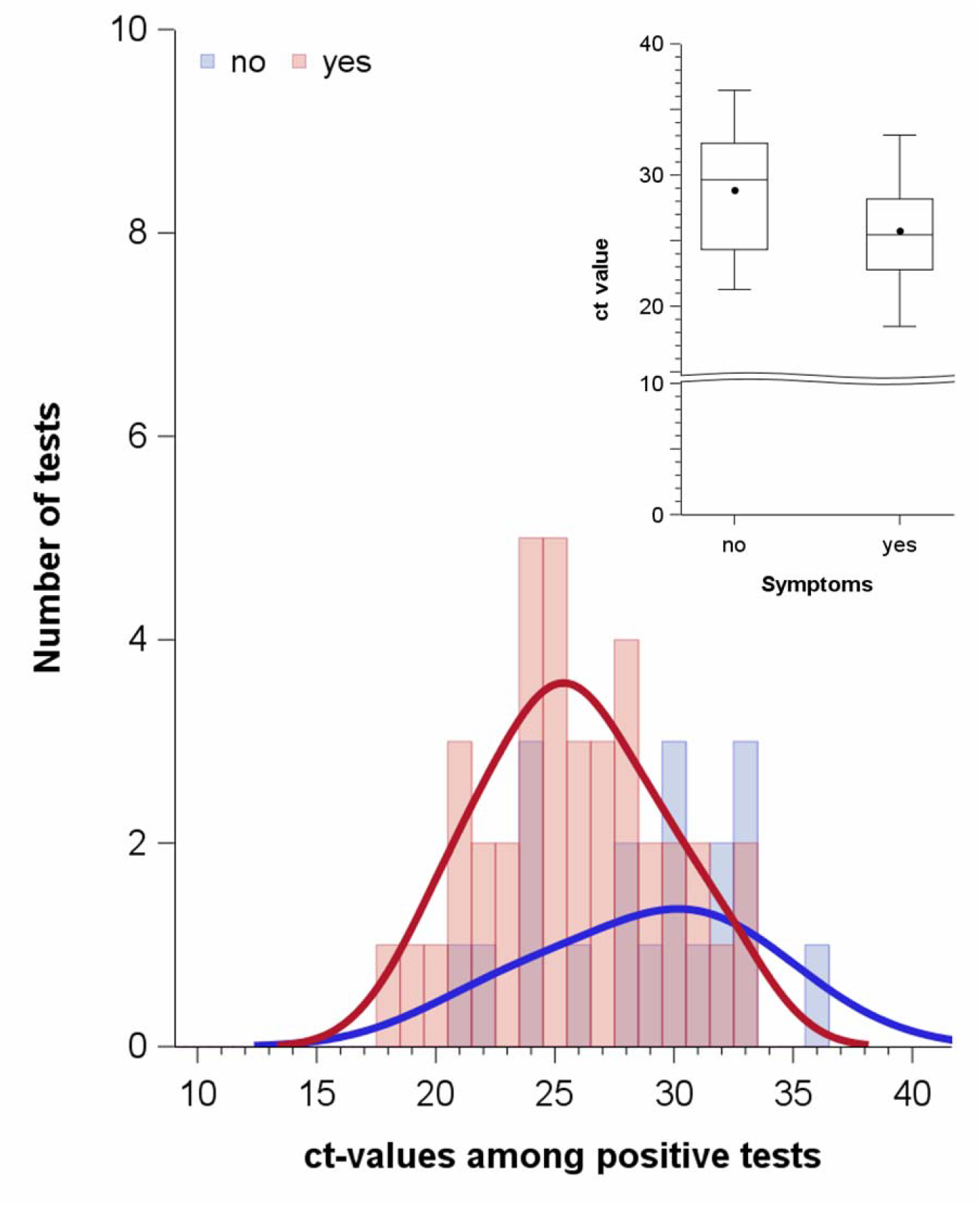
Ct value distribution among symptomatic and asymptomatic individuals’ with positive tests in the region of Münster, North Rhine-Westphalia, Germany. Legend: “no” means “no symptoms”, “yes” means “symptoms”; dots in the box plot indicate mean values and horizontal lines in the boxes indicate median values. Asymptomatic individuals : n=19, median 29.6, mean 28.8, SD 4.3; symptomatic individuals: n=39 median 25.5, mean 25.8, SD 3.7

The percentage of Ct values below 25 was lower among children (0-9) and adolescents (10-19), as well as among older age groups, 70-79, 80-89, 90-99 years (21.0%, 38.2%, and 35.3%, 37.4 %, 37.0 %, respectively) compared to people aged 20-69 years (43.4%) (**Figure 2**).

**Figure 2.**
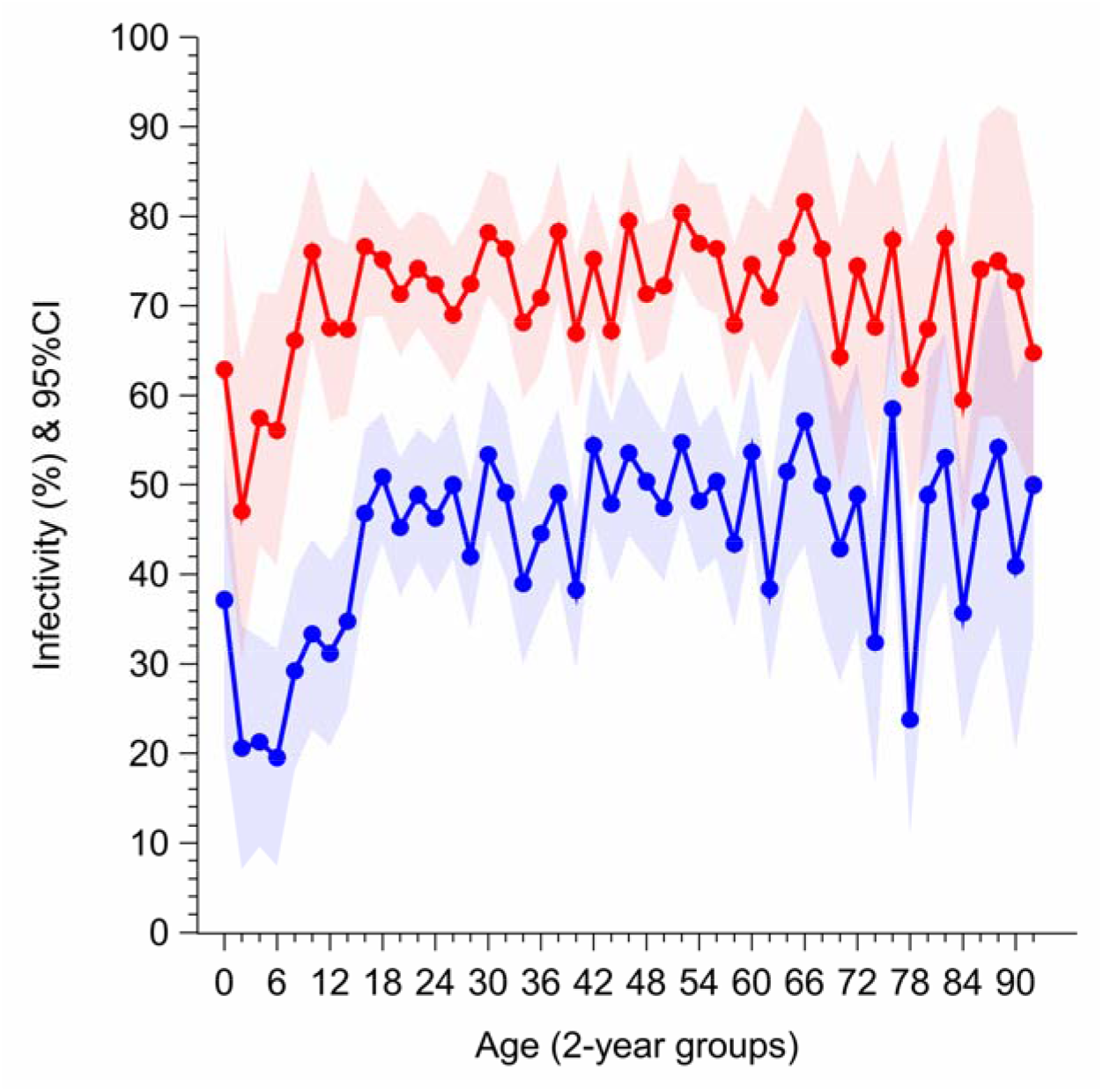
Percentages of SARS-CoV-2 infected people with Ct values below 25 and below 30 in the region of Münster, North Rhine-Westphalia, Germany. Legend: percentage of SARS-CoV-2 infected people with Ct value cutoff < 25 (blue graph); percentage of SARS-CoV-2 infected people with Ct value cutoff < 30 (red graph).

A comparison of Ct value distributions among positively tested subjects in the population-based samples of Münster and the ONS COVID-19 Infection Survey shows that median Ct values were higher (29.8 and 27.7) in Münster for the first two weeks in October, thereafter declining (24.4, 25.4) and staying at a lower level for the remaining weeks compared to the UK. In contrast to Münster, the weekly median Ct values in the UK steadily increased from 25.5 (first week in October) to 30.6 (first week in December) (**Table 2**).

**Table 2.** Distribution of Ct values among positively tested people for the population of Münster, Germany and United Kingdom for identical time periods in 2020.

| Characteristic | N | 10th Pctl | 25th Pctl | Median | 75th Pctl | 90th Pctl | Mean |
| --- | --- | --- | --- | --- | --- | --- | --- |
| Data from Münster, Germany |  |  |  |  |  |  |  |
| 30.09.-06.10 | 177 | 20.9 | 23.7 | 29.8 | 32.8 | 34.6 | 28.4 |
| 07.10.-13.10. | 282 | 19.5 | 22.1 | 27.7 | 31.3 | 32.9 | 26.8 |
| 14.10.-20.10. | 266 | 18.2 | 20.9 | 24.4 | 28.8 | 31.7 | 24.8 |
| 21.10.-28.10. | 473 | 18.9 | 21.5 | 25.4 | 29.6 | 32.4 | 25.6 |
| 29.10.-03.11. | 438 | 20.3 | 22.6 | 26.7 | 30.7 | 33.2 | 26.6 |
| 04.11.-10.11. | 399 | 20.2 | 23.0 | 27.2 | 31.2 | 33.6 | 27.1 |
| 11.11.-17.11. | 330 | 20.1 | 22.5 | 26.6 | 30.5 | 32.8 | 26.5 |
| 18.11.-24.11. | 391 | 19.0 | 21.7 | 26.2 | 30.3 | 33.4 | 26.0 |
| 25.11.-01.12. | 342 | 19.5 | 22.1 | 26.8 | 30.9 | 33.4 | 26.5 |
| 02.12.-09.12. | 194 | 19.5 | 22.9 | 25.9 | 30.3 | 32.3 | 26.1 |
| Data from United Kingdom |  |  |  |  |  |  |  |
| 30.09.-06.10 |  | 16.1 | 19.4 | 25.5 | 30.7 | 33.3 | 24.9 |
| 07.10.-13.10. |  | 16.6 | 20.1 | 25.7 | 31.3 | 33.6 | 25.5 |
| 14.10.-20.10. |  | 17.3 | 21.3 | 27.5 | 31.9 | 33.8 | 26.4 |
| 21.10.-28.10. |  | 17.0 | 20.9 | 27.2 | 31.7 | 33.6 | 26.1 |
| 29.10.-03.11. |  | 16.8 | 20.9 | 27.3 | 31.7 | 33.6 | 26.2 |
| 04.11.-10.11. |  | 17.0 | 21.6 | 28.5 | 31.8 | 33.6 | 26.6 |
| 11.11.-17.11. |  | 17.0 | 21.3 | 29.1 | 32.5 | 34.0 | 27.0 |
| 18.11.-24.11. |  | 18.5 | 24.0 | 29.8 | 32.8 | 34.2 | 28.0 |
| 25.11.-01.12. |  | 18.3 | 24.9 | 30.9 | 33.6 | 34.6 | 28.7 |
| 02.12.-09.12. |  | 17.4 | 23.0 | 30.6 | 33.5 | 34.6 | 28.0 |

Legend Table 2: Office for National Statistics, COVID-19 Infection Survey; Ct values for all genes (date of publication Dec 21, 2020)

Statistical analyses on a case basis that includes repetitive testing, rather than on a person basis did not reveal relevant differences (**Supplementary Table 2, Supplementary Figures 3-6**).

## Discussion

We found that the RT-PCR test positive rate heavily depends on the national testing strategy. The positive rate, furthermore depends on age, with lower rates among young children and the elderly. The majority of positive tests in our sample showed Ct values of 25 or higher, indicating a low viral load. Among positively tested children (0-9 years) and elderly (≥ 70 years) higher Ct values (>25) were recorded even more frequently. Ct values were on average lower among symptomatic compared to asymptomatic subjects. The distribution of Ct values over time did not show great variability with median Ct-values mostly above 25 in all calendar weeks.

Our study has two main strengths. First, we used a single RT-PCR platform for all tests. Second, we provide population-based data on test positivity and Ct values over a period of 39 weeks in 2020. However, there are several factors that limit our results. First, all our Ct values were obtained with a single assay limiting the comparability of our results to those obtained with other PCR test platforms and assays. Second, we were able to classify RT-PCR test positive individuals as asymptomatic or symptomatic only for a very small fraction of our sample. There is evidence that the degree of symptoms among SARS-CoV-2 infected people is inversely associated with the Ct value [10]. Third, even though the missing values mechanism indicates that bias was unlikely, it must be noted that we were unable to retrieve Ct values for a period of several weeks extending from the end of April through June.

Our results are similar to the observations made in the ONS Survey with consistenly low positive rates (0.06%) during the summer months, followed by a rise to more than 1% by the end of October 2020. A substantial proportion (45%-68%) of test positive individuals in the UK did not report symptoms around their positive test [11]. To date the prevalence of SARS-CoV-2 infection in the German population is unknown except for estimates in hot spots ranging from a seroprevalence of 14.7% in Gangelt (March 2020), 6% and 8 % in Bad Feilenbach and Künzelsau, respectively, in summer of 2020 and 4.4% in Berlin-Mitte from November to December 2020 [12]. Data on the association between positive RT-PCR tests and the presence of sero-antibodies against SARS-CoV-2 are sparse, calculation of the pretest probability and positive predictive value of a positive RT-PCR screening test remains hypothetical. Testing predominantly symptomatic individuals in our sample increased the percentage of positive tests, however, this might also be related to other coronaviruses causing flu like symptoms being detected by our test. The analytical accuracy of the SARS-CoV-2 RT-PCR test was only validated in a limited number of laboratory specimens of known respiratory pathogens. The diagnostic accuracy, i.e. the ability to reliably separate people with SARS-CoV-2 infection from those without such infection including the probability of false positive and false negative results was never assessed in the population. Particularly, in the asymptomatic population during the summer months, with an assumed very low pretest probability (50 per 100,000) the positive predictive value of a positive RT-PCR test was calculated to have been as low as 0.7%. [13].

According to the Robert Koch-Institute (RKI) and the WHO, a positive RT-PCR test defines a SARS-CoV-2 infected case; these cases are accumulated to weekly incidence estimates. If incidences of 35 or 50 cases per 100,000 per week occur, the national pandemic legislation entitles local and state governments to impose non-pharmaceutical intervention (NPI) measures. In light of our findings that more than half of these cases are not likely to be infectious, RT-PCR test positivity should not be taken as an accurate measure of infectious SARS-CoV-2 incidence. Real world RT-PCR positive test results most likely comprise a mixture of prevalent cases (contact with SARS-CoV-2 at least 9 days ago, most likely not infectious), new cases (contact with virus less than 9 days, more likely to be infectious) and false positives. In addition, the clinical relevance of a positive RT-PCR test during the different time periods from March to December 2020 is also unclear [14]. From an epidemiological point of view it is similarly problematic to relate COVID-19 deaths to a cumulative incidence based on RT-PCR test positives, rather than from estimates based on data obtained from sero-prevalence studies [15].

We compared the distribution of Ct values between calendar weeks 38-49 to the respective ONS survey data, and found a similar range of median Ct values (24.4 to 29.8 in Münster and 25.5 to 30.9 in the UK); of note, both surveys revealed that more than half of the Ct value distribution was above 25, except for calendar week 39 in Münster. However, the direct comparison of the Ct value distributions of the two surveys may be limited by differences in the design of the assay.

It has been repeatedly reported that children appear to be less frequently infected by SARS-CoV-2 than adults [16, 17] and, when infected, typically have mild symptoms [17, 18]. Furthermore, there is evidence that a higher viral load is associated with more severe clinical courses of COVID-19 [19, 20]. Among RT-PCR-test positive persons, we found that children and adolescents had a lower percentage of high viral load, corroborating findings of no transmission of COVID-19 from children and adolescents [21]. In contrast, among 145 positively tested hospitalized patients with moderate illness within one week of symptoms in Chicago, children younger than 5 years had markedly lower Ct values (Abbot RealTime SARS-CoV-2 assay on the m2000 RealTime System, n=46, median 6.5) than children aged 5-17 years (n=51, 11.1) and adults aged 18-65 years (n=48, 11.0) [22]. Again, methodological differences between RT-PCR platforms limit the interpretation of results. Severe SARS-CoV-2 infections among children are rare. For example, from March through June 2020, overall only 15 children aged 1-16 years in Sweden with COVID-19 were admitted to an intensive care unit, which equals 1 child in 130,000 [23].

Our results corroborate findings that assessment and control of infectiousness by routine reporting of the number of “positive” RT-PCR tests, as a gold standard, ignores “that 50-75% of the time an individual is PCR positive, they are likely to be post-infectious” [14]. Asymptomatic RT-PCR test positive individuals have higher Ct values and a lower probability of being infective than test positives with symptoms. Although Ct values have been shown to be inversely associated with viral load and infectivity, there is no international standardization across laboratories, making the interpretation of RT-PCR tests as a tool for mass screening confusing/difficult. In conclusion, our results challenge the current policy of relying on RT-PCR test screening as a base for detecting SARS-CoV-2 infection in the population and for pandemic decision making, including NPI measures such as quarantine, isolation, and lockdown.

## Data Availability

Anonymised data are available on reasonable request and can be provided by the corresponding author.

