## Supplementary File for "The performance of the SARS-CoV-2 RT-PCR test as a tool for detecting SARS-CoV-2 infection in the population: A survey of routine laboratory RT-PCR test results from the region of Münster, Germany" (from another version of this preprint: version 1, 2021-05-11)

**S U P P L E M E N T**

**Suppl. Table 1	Distribution of Ct values among positively tested people in the region of Münster, North Rhine-Westphalia, Germany**

| **Characteristic** | **N** | **Minimum** | **10th Pctl** | **25th Pctl** | **Median** | **75th Pctl** | **90th Pctl** | **Maximum** | **Mean** | **SD** |
| --- | --- | --- | --- | --- | --- | --- | --- | --- | --- | --- |
| Overall | 3810 | 13.02 | 19.49 | 22.23 | 26.69 | 30.77 | 33.19 | 38.58 | 26.51 | 5.17 |
| **Sex** |  |  |  |  |  |  |  |  |  |  |
| Unknown | 18 | 18.47 | 19.71 | 23.80 | 28.79 | 32.08 | 34.08 | 35.11 | 27.39 | 5.23 |
| Male | 1815 | 13.60 | 19.40 | 21.93 | 26.57 | 30.85 | 33.13 | 37.64 | 26.40 | 5.26 |
| Female | 1977 | 13.02 | 19.62 | 22.59 | 26.75 | 30.72 | 33.20 | 38.58 | 26.61 | 5.10 |
| **Age (years)** |  |  |  |  |  |  |  |  |  |  |
| unknown | 17 | 18.21 | 23.17 | 29.28 | 32.65 | 35.06 | 35.52 | 36.09 | 31.29 | 4.91 |
| 0-9 | 209 | 15.70 | 21.43 | 25.87 | 29.42 | 31.84 | 34.14 | 37.54 | 28.57 | 4.67 |
| 10-19 | 503 | 13.02 | 19.69 | 23.23 | 27.16 | 30.59 | 32.80 | 38.00 | 26.77 | 4.86 |
| 20-29 | 682 | 15.18 | 19.47 | 22.08 | 26.42 | 30.72 | 32.81 | 38.58 | 26.35 | 5.14 |
| 30-39 | 527 | 15.53 | 19.85 | 22.20 | 26.04 | 30.45 | 33.34 | 37.32 | 26.33 | 5.07 |
| 40-49 | 544 | 14.19 | 19.38 | 21.59 | 26.33 | 30.99 | 33.25 | 37.15 | 26.27 | 5.38 |
| 50-59 | 608 | 13.60 | 19.08 | 21.50 | 25.94 | 30.35 | 33.15 | 36.87 | 25.98 | 5.26 |
| 60-69 | 328 | 14.99 | 19.27 | 21.89 | 25.56 | 30.23 | 32.77 | 37.64 | 25.97 | 5.15 |
| 70-79 | 190 | 14.43 | 19.96 | 22.59 | 27.70 | 31.25 | 33.82 | 36.73 | 27.13 | 5.21 |
| 80-89 | 155 | 14.41 | 19.32 | 22.92 | 27.89 | 30.90 | 32.75 | 36.80 | 26.82 | 5.24 |
| 90-99 | 46 | 15.87 | 17.28 | 23.25 | 27.99 | 31.45 | 33.51 | 35.00 | 26.97 | 5.43 |
| Calendar week |  |  |  |  |  |  |  |  |  |  |
| 10-19 | 77 | 16.62 | 20.26 | 25.15 | 30.36 | 32.73 | 34.45 | 36.00 | 28.74 | 5.10 |
| 20-44 | 2292 | 13.02 | 19.50 | 22.25 | 26.72 | 30.73 | 33.22 | 38.58 | 26.52 | 5.20 |
| 45-49 | 1441 | 13.60 | 19.45 | 22.12 | 26.52 | 30.70 | 32.94 | 37.21 | 26.38 | 5.12 |

**Suppl. Table 2	Characteristics of PCR tests in the region of Münster, North	Rhine-Westphalia, Germany, March 26 - December 6, 2020**

|  | **Number of tests^1)^** | **Positive tests** | | **Mean Ct value among positive tests^2)^** | | **Percentage of positive tests with Ct values ^2)^** | |
| --- | --- | --- | --- | --- | --- | --- | --- |
|  | **N** | **N** | **%** | **Mean** | **SD** | **< 25** | **<35** |
| All | 193258 |  |  | 26.8 | 5.2 | 38.6 | 67.2 |
| Men | 80516 | 2308 | 2.9 | 26.7 | 5.3 | 39.5 | 67.2 |
| Women | 112436 | 2659 | 2.4 | 26.9 | 5.2 | 37.8 | 67.2 |
| Unknown | 306 | 18 | 5.9 | 27.4 | 5.2 | 38.9 | 66.7 |
| Swab site |  |  |  |  |  |  |  |
| Nose & throat | 11013 | 310 | 2.8 | 27.0 | 5.4 | 34.6 | 65.5 |
| Throat | 8294 | 171 | 2.1 | 26.6 | 4.7 | 39.0 | 74.0 |
| Unspecified/other | 173951 | 4504 | 2.6 | 26.8 | 5.2 | 38.9 | 67.1 |
| Age group |  |  |  |  |  |  |  |
| 0-9 | 10457 | 244 | 2.3 | 28.5 | 4.6 | 21.3 | 57.0 |
| 10-19 | 16296 | 578 | 3.6 | 26.7 | 4.9 | 38.1 | 71.6 |
| 20-29 | 24851 | 846 | 3.4 | 26.4 | 5.2 | 41.1 | 69.0 |
| 30-39 | 25442 | 640 | 2.5 | 26.5 | 5.1 | 41.9 | 71.0 |
| 40-49 | 25313 | 686 | 2.7 | 26.5 | 5.5 | 42.5 | 67.4 |
| 50-59 | 30457 | 780 | 2.6 | 26.7 | 5.3 | 41.4 | 68.5 |
| 60-69 | 21181 | 435 | 2.1 | 27.7 | 5.1 | 40.1 | 67.3 |
| 70-79 | 15440 | 330 | 2.1 | 27.6 | 5.1 | 30.6 | 60.6 |
| 80-89 | 18075 | 331 | 1.8 | 26.7 | 5.4 | 32.4 | 59.9 |
| 90-99 | 5146 | 96 | 1.9 | 26.7 | 5.4 | 37.8 | 65.9 |
| 100+ | 40 | 1 |  |  |  |  |  |
| unknown | 560 | 18 | 3.2 | 31.3 | 4.9 | 11.8 | 29.4 |
| Calendar week |  |  |  |  |  |  |  |
| 10-19 | 14348 | 361 | 2.5 | 28.7 | 5.0 | 21.7 | 48.2 |
| 20-44 | 157151 | 2746 | 1.8 | 26.7 | 5.3 | 39.3 | 67.7 |
| 45-49 | 21759 | 1878 | 8.6 | 26.8 | 5.2 | 38.5 | 67.3 |
| Specific phases of the pandemic^3)^ |  |  |  |  |  |  |  |
| Peak 1^st^ wave | 2228 | 36 | 1.6 | 27.8 | 5.4 | 26.5 | 55.9 |
| Traveler return | 19079 | 73 | 0.4 | 28.9 | 5.4 | 26.4 | 55.6 |
| Peak 2^nd^ wave | 5161 | 516 | 10.0 | 27.2 | 5.0 | 34.1 | 65.5 |

Legend table: SD = standard deviation; ^1)^ only persons with tests that were clearly either positive or negative were included; ^2)^ among 4985 positive tests, the Ct value was available for 4559 tests (91.5%); Ct values were not retrievable for positive tests during the calendar weeks 12-13 and 16-25 in 2020; standard deviations do not account for repeated measurements; ^3)^ Peak of 1^st^ wave in weeks 12-13 (16.-29.3.2020); proxy weeks 13-14; unselective testing in weeks 33-34 (peak of tests for traveler return); peak of 2^nd^ wave in weeks 50-51 (7.-20.12.2020), proxy weeks 48-49

**Supplementary Figure 1**

**Number of weekly PCR tests in 2020 and percentage of positive tests including 95% confidence intervals in the region of Münster, North Rhine-Westphalia, Germany**

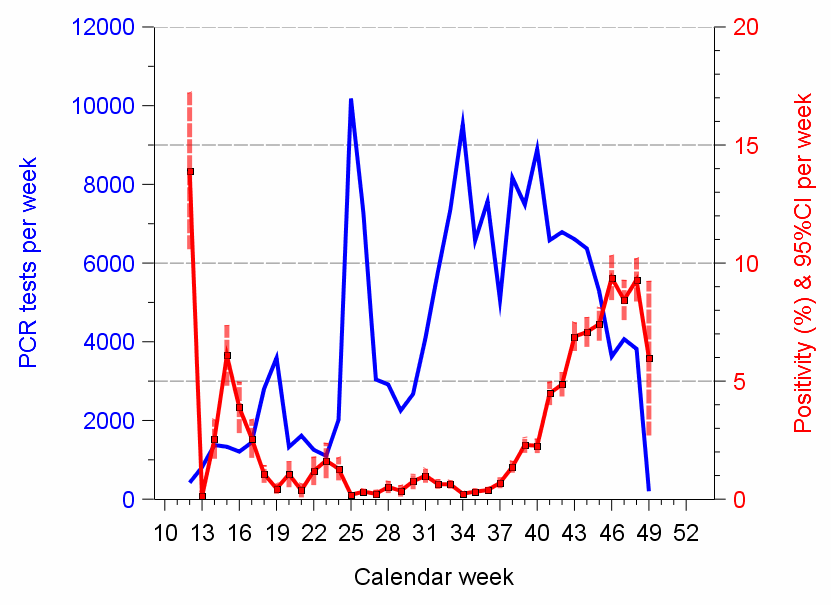

Legend: vertical dotted red lines indicate pointwise 95% confidence intervals

**Supplementary Figure 2**

**Number of weekly positive PCR tests in 2020 and mean Ct values including 95% confidence intervals in the region of Münster, North Rhine-Westphalia, Germany**

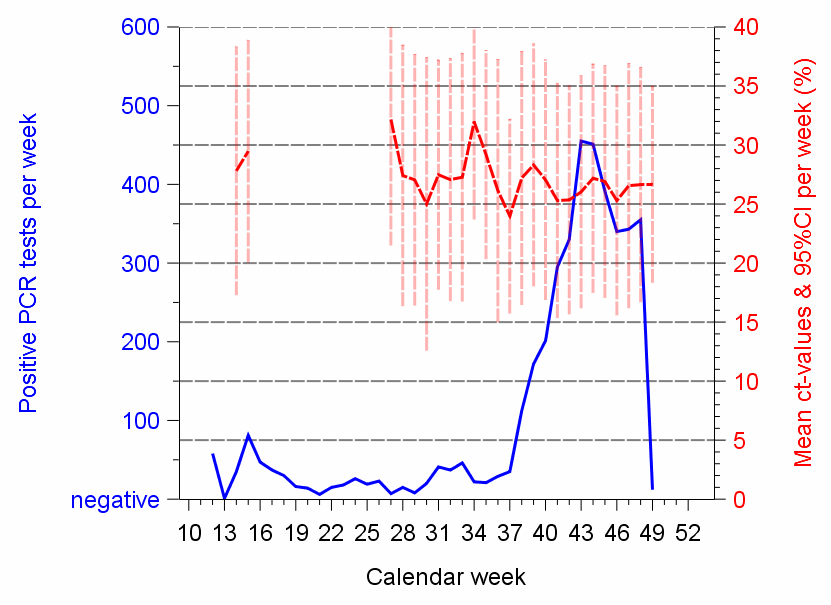

Legend: vertical dotted red lines indicate pointwise 95% confidence intervals

**Suppl. Figure 3**

**Number of weekly PCR tests in 2020 and percentage of positive tests including 95% confidence intervals in the region of Münster, North Rhine-Westphalia, Germany**

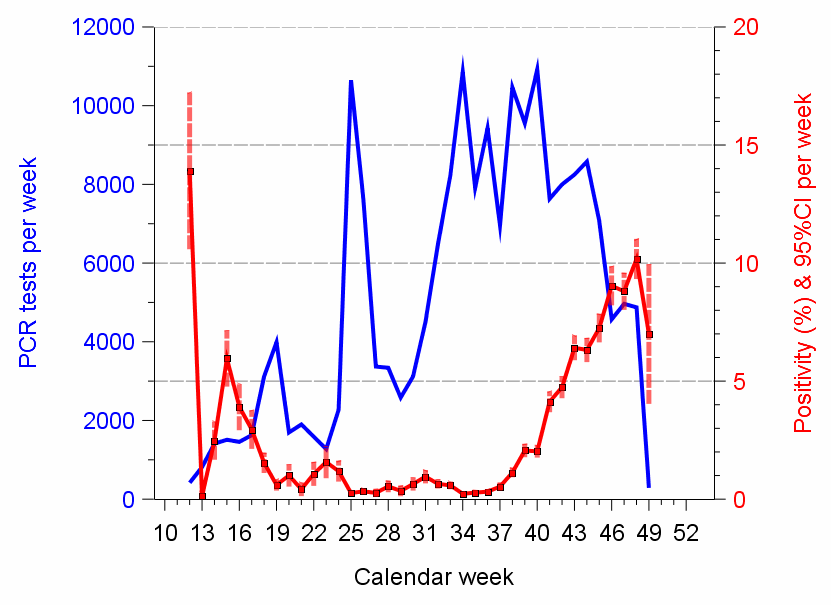

Legend: vertical dotted red lines indicate pointwise 95% confidence intervals; confidence intervals do not account for repeated measurements;

**Suppl. Figure 4**

**Number of weekly positive PCR tests in 2020 and mean Ct values including 95% confidence intervals in the region of Münster, North Rhine-Westphalia, Germany**

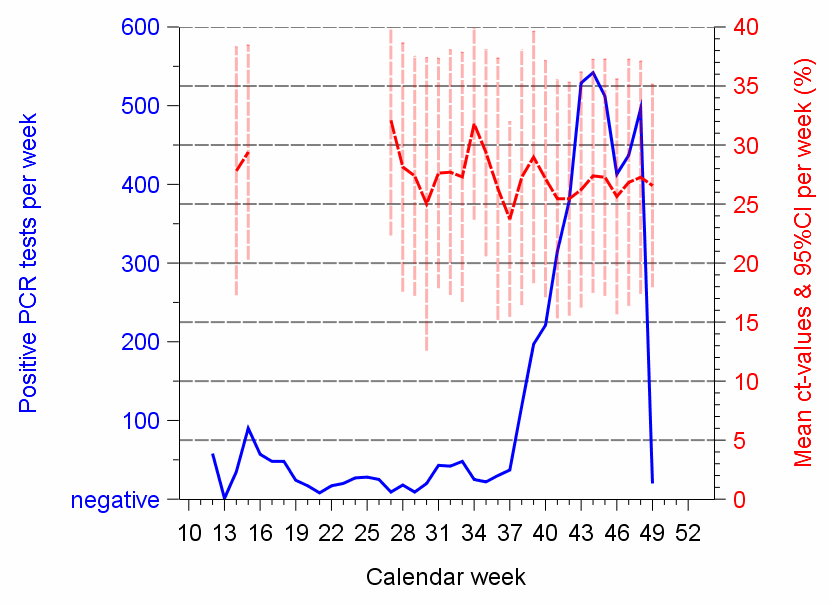

Legend: vertical dotted red lines indicate pointwise 95% confidence intervals; confidence intervals do not account for repeated measurements;

**Suppl. Figure 5**

**Ct value distribution of symptomatic and asymptomatic patients with positive tests in the region of Münster, North Rhine-Westphalia, Germany**

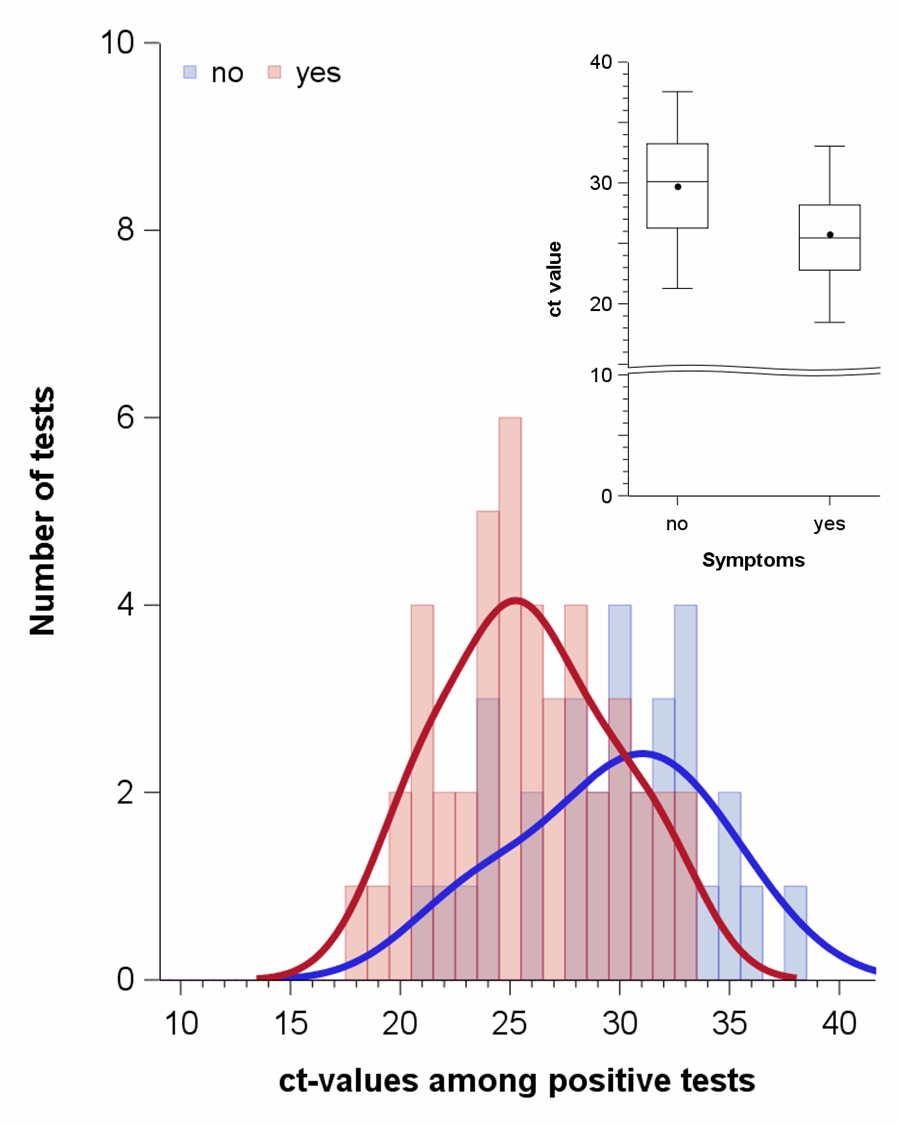

Legend: “no” means “no symptoms”, “yes” means “symptoms”; dots in the box plot indicate mean values and horizontal lines in the boxes indicate median values; tests among asymptomatic patients: n=31, median 30.1, mean 29.7, SD 4.3; tests among symptomatic patients: n=45, median 25.5, mean 25.8, SD 3.8

**Suppl. Figure 6**

**Percentages of SARS-CoV-2 infected people with Ct values below 25 and below 30 in the region of Münster, North Rhine-Westphalia, Germany**

Legend: percentage of SARS-CoV-2 infected people with Ct value cutoff < 25 (blue graph); percentage of SARS-CoV-2 infected people with Ct value cutoff < 30 (red graph).
